# CPAP/BiPAP Compliance Improves Survival in LVAD Recipients with Obstructive Sleep Apnea

**DOI:** 10.64898/2026.04.20.26351345

**Authors:** Jason Carlquist, Shane S. Scott, Jonathan C. Wright, Ma Jianing, Jing Peng, Nahush A. Mokadam, Bryan A. Whitson, Sakima A. Smith

## Abstract

**Purpose:** Obstructive sleep apnea (OSA) is a common comorbidity in heart failure (HF) patients with prevalence increasing as HF severity worsens. While CPAP/BiPAP has been shown to reduce disease burden and mortality in the general HF population, it is unclear whether these benefits extend to patients with left ventricular assist devices (LVADs). We sought to determine whether OSA affects long-term survival in newly implanted LVAD patients and whether CPAP/BiPAP treatment confers mortality benefits.

**Methods:** This single-center retrospective study included patients who underwent LVAD implantation between January 2007 and February 2022. Recipients were stratified by OSA status (OSA vs No-OSA), and those with OSA were further categorized based on CPAP/BiPAP compliance. Comparative statistics and Kaplan–Meier survival analyses were performed, with log-rank tests used to compare groups and assess survival differences. A Cox proportional hazards model was conducted to evaluate the association between risk factors and survival among patients with OSA and No-OSA.

**Results:** Before LVAD implantation, patients with OSA had higher body mass index, hypertension, and a higher rate of implantable cardioverter-defibrillator placement than those without OSA. OSA was not associated with increased postoperative complications. Although survival did not differ significantly between OSA and No-OSA patients (p=0.33), CPAP/BiPAP-compliant OSA patients had significantly better survival than noncompliant patients (p=0.0099).

**Conclusions:** LVAD patients with OSA who consistently use CPAP/BiPAP have better survival than those who do not. CPAP/BiPAP is a simple, low-risk treatment that can reduce mortality in this population. Therefore, increased perioperative screening for OSA should be considered for patients receiving LVADs. Multicenter studies are needed to confirm our findings further.

## INTRODUCTION

Obstructive sleep apnea (OSA) is a common comorbidity in patients with heart failure (HF), and its prevalence increases with HF severity (1). Compared to HF patients without OSA, those with untreated OSA have higher morbidity and mortality (2). Mechanistically, episodes of apnea and hypopnea result in intermittent hypoxemia, which subsequently increases heart rate, blood pressure, afterload, and myocardial oxygen demand. These changes are associated with the development and progression of heart failure. In addition, the negative intrathoracic pressures generated during apneic episodes reduces cardiac output, worsening systolic and diastolic function in HF patients.

Mechanical support devices, including left ventricular assist devices (LVADs), have improved survival in advanced HF. However, despite OSA affecting 20-60% of HF patients (4), its impact on those with LVADs remains unclear. Given the unique physiology of LVAD-supported hearts, understanding the interplay between OSA and LVADs is important.

Prior studies suggest that OSA precludes and/or contributes to HF development and worse outcomes (4). Kumai et al. found that sleep-disordered breathing, including OSA and central sleep apnea (CSA), was prevalent in 24% of LVAD patients, with OSA linked with to more frequent ventricular tachyarrhythmias compared to the No-OSA patients (6). Interestingly, LVAD implantation can also significantly influence the severity of sleep-disordered breathing, with some cases showing complete resolution of CSA and/or OSA (7–8) and improvements in patient-reported HF symptoms and sleepiness (7). Following heart transplantation, however, both the CSA and OSA recurred (8).

Noninvasive positive pressure ventilation (NIPPV), including positive airway pressure (CPAP) and bilevel positive airway pressure (BiPAP), is the standard treatment for OSA. CPAP/BiPAP use has been associated with reduced mortality, fewer arrhythmias, and reductions blood pressure in patients (9). Several single-center studies have reported benefits associated with CPAP use in HF patients, including improved left ventricular function, reduced sympathetic tone and myocardial oxygen consumption, and lower rates of HF hospitalization and mortality (2,10–12). The multicenter Sleep Apnea cardioVascular Endpoints (SAVE) trial assessed 2717 patients with moderate to severe OSA and established cardiovascular disease and found no reduction in major cardiovascular events among OSA patients treated with CPAP (13); however, patients with overt HF or LVADs were excluded.

Hence, we hypothesized that LVAD patients with OSA have worse outcomes than those without OSA, and CPAP/BiPAP compliance may improve survival and reduce disease burden in this population.

## METHODS

### Ethics and IRB Approval

This study was conducted in accordance with the rules and principles set forth by the Institutional Review Board at The Ohio State University. The study was reviewed and approved by the Institutional Review Board (IRB Approval Number: 2019H0328).

### Definition of the Variables

| Variable | Definition |
| --- | --- |
| **OSA (Obstructive Sleep Apnea)** | Disease characterized by repetitive collapse of the upper airway during sleep, which culminates in periods of apnea (complete airflow cessation) and hypopnea (reduced airflow) due to attempts at breathing against an occluded airway. |
| **AHI (Apnea Hypopnea Index)** | Total number of apneas and hypopneas per hour of sleep. One common index used to diagnose and classify the severity of OSA. |
| **Mild OSA** | AHI between 5 and 14 events per hour. |
| **Moderate OSA** | AHI between 15 and 30 events per hour. |
| **Severe OSA** | AHI greater than 30 events per hour. |
| **RVF (Right ventricular failure)** | RVF was defined as either 1) need for short- or long-term right-sided circulatory support, 2) continuous inotropic support for >_14 days, or 3) inhaled nitric oxide ventilation for >48 h post-operatively, as in the original EUROMACS-RHF derivation study (14). |

### Study Population

This retrospective single-center study included 514 patients who underwent LVAD implantation between January 2007 and February 2022 at The Ohio State University Wexner Medical Center. Patients with an ICD-9 or ICD-10 (G47.33) diagnosis of OSA were identified, while those with CSA was excluded. Inclusion criteria required a confirmed OSA diagnosis by polysomnography (PSG) prior to LVAD implantation. Patients with chart-documented OSA but no PSG confirmation were excluded. Among those with PSG-confirmed OSA, severity classified by apnea-hypopnea index (AHI). Compliance was defined pragmatically based on provider documentation of consistent use at clinic follow-ups. Only patients with documented compliance or noncompliance with CPAP/BiPAP prior to LVAD implantation were included; those who became noncompliant after implantation were excluded (**Figure 1**).

**Figure 1.**
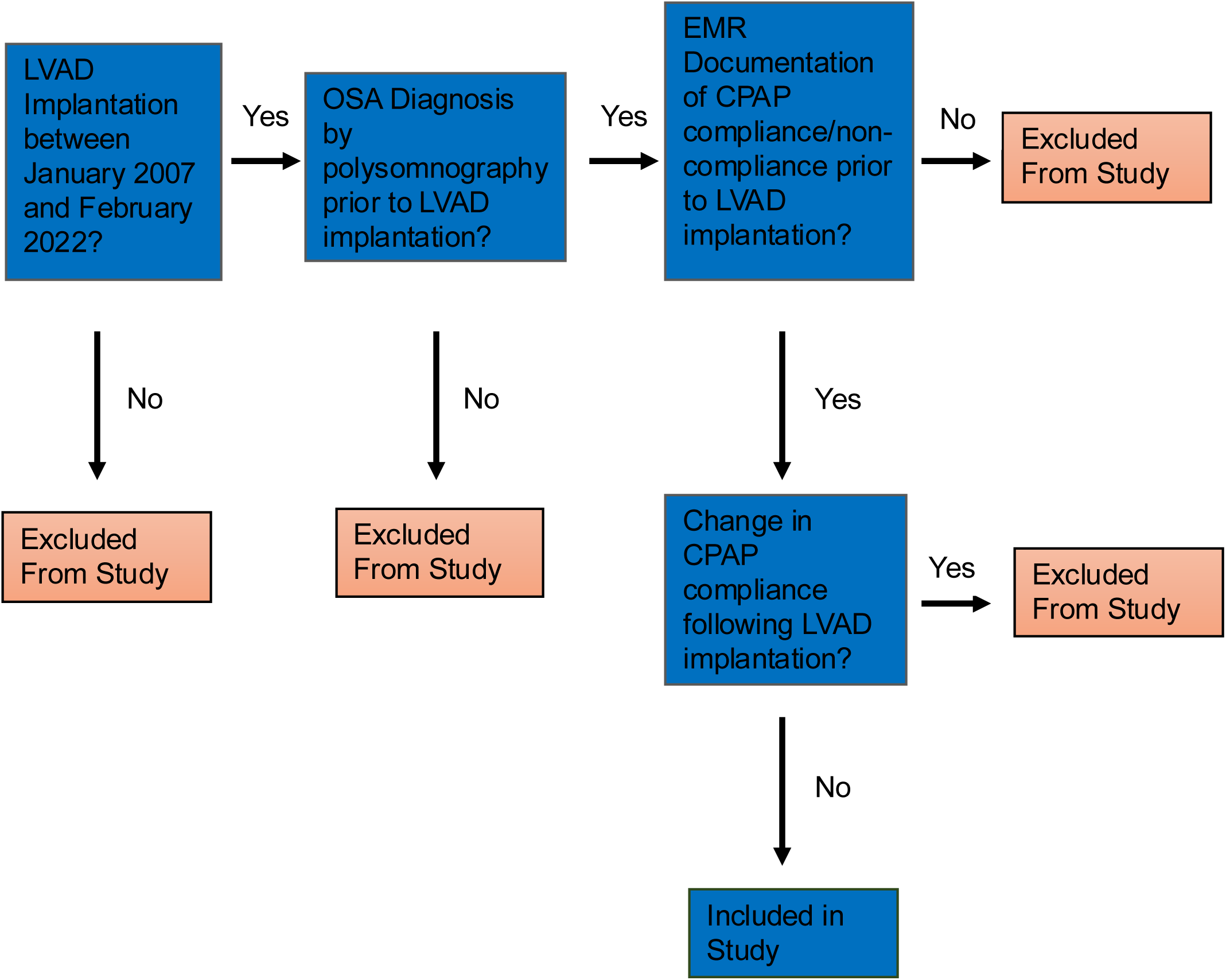
Consort Diagram for Study Population. Visual description of inclusion and exclusion criteria required for LVAD patients with OSA participation in the study.

### Data Analysis

The primary outcome was all-cause mortality. Time-to-event analysis was used to evaluate survival differences based on OSA status, OSA severity, and CPAP/BiPAP compliance among LVAD patients. Multivariate analysis was performed to adjust for baseline demographics, comorbidities, and use of medical or surgical therapies.

### Statistical Analysis

Demographic and clinical variables of LVAD patients with OSA were compared to those without OSA. Normally distributed continuous variables were summarized as mean ± standard deviation, and skewed variables as median with interquartile range. Categorical variables were summarized as frequencies and percentages. Kaplan–Meier curves and log-rank tests were used to compare overall survival after LVAD implantation between OSA and No-OSA patients. The start time was defined as the date of OSA diagnosis, and the end time as the date of death or December 31, 2022, for uncensored patients. Cox proportional hazards models were used to evaluate differences in survival between groups, adjusting for relevant prognostic factors. Among OSA patients, survival was also compared by OSA severity (mild, moderate, severe) and CPAP/BiPAP compliance. P-values were adjusted for multiple comparisons using the false discovery rate (FDR) method. An adjusted p value < 0.05 was considered statistically significant. All statistical analysis was done using R software, version 4.5.0.

## RESULTS

### Baseline Characteristics Pre-LVAD Implantation

A total of 514 LVAD patients were identified, of whom 468 (91.1%) patients had No-OSA, and 46 (8.9%) had OSA. Among the recipients, there were no significant differences in age, sex, or LVAD type between the OSA and No-OSA groups (adjusted p > 0.05).

Prior to LVAD implantation, patients with OSA had significantly higher body mass index (31.48 ± 6.39 vs. 27.04 ± 5.93; p < 0.001; adjusted p = 0.012). However, some comorbidities and baseline medications differed between groups (**Table 1**). OSA patients were more likely to have hypertension (p = 0.002; adjusted *p* = 0.02) and had higher rates of implantable cardioverter defibrillator (ICD) placement (p=0.004; adjusted p = 0.033). They were also more frequently prescribed angiotensin-aldosterone inhibitor therapy, including aldosterone antagonists, angiotensin receptor blockers (ARBs), and beta-blockers (all adjusted p ≤ 0.05; **Table 1**). No significant differences were observed when comparing the prevalence of diabetes, COPD, CAD, prior MI, stroke, or inotropes use.

**Table 1:**
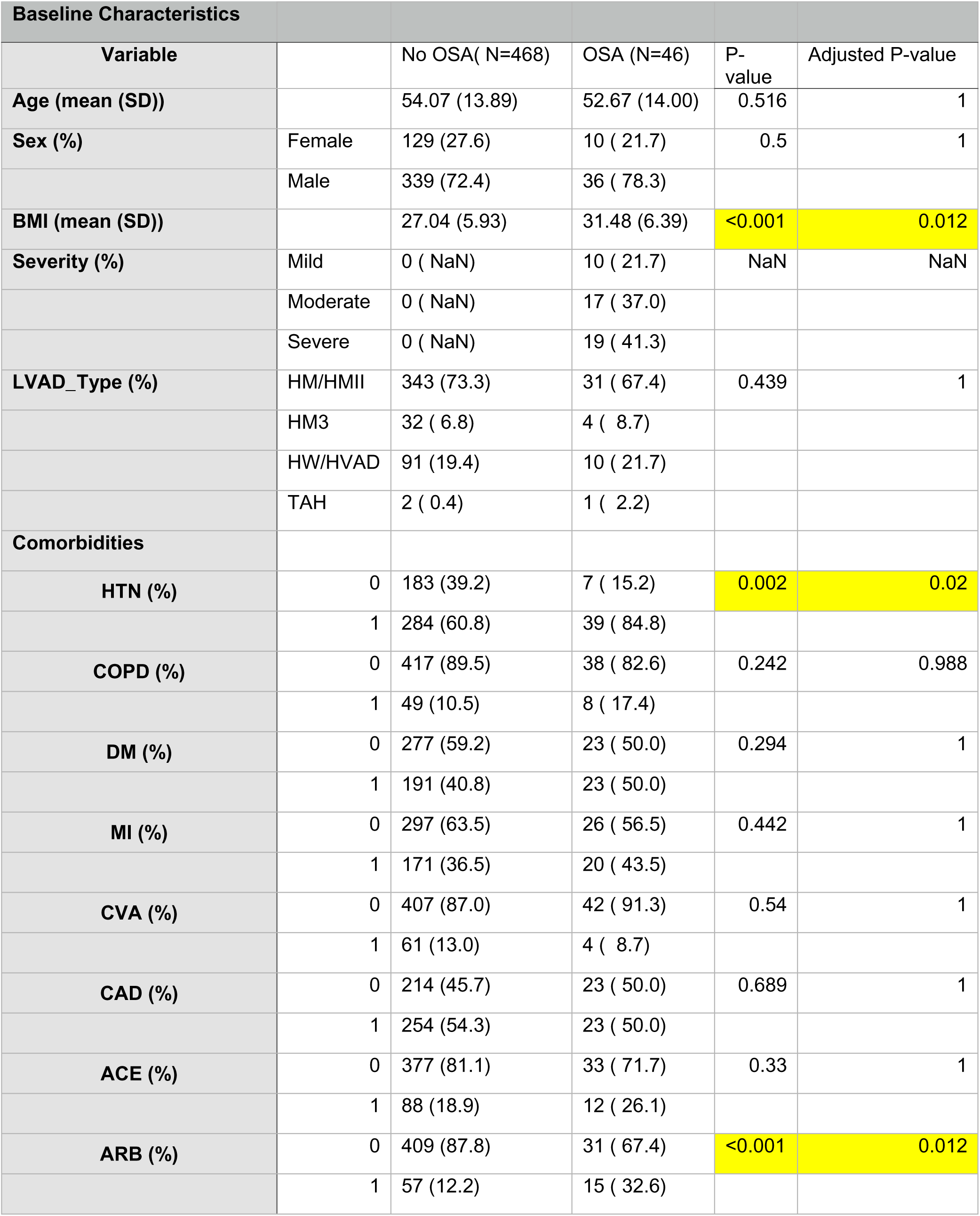
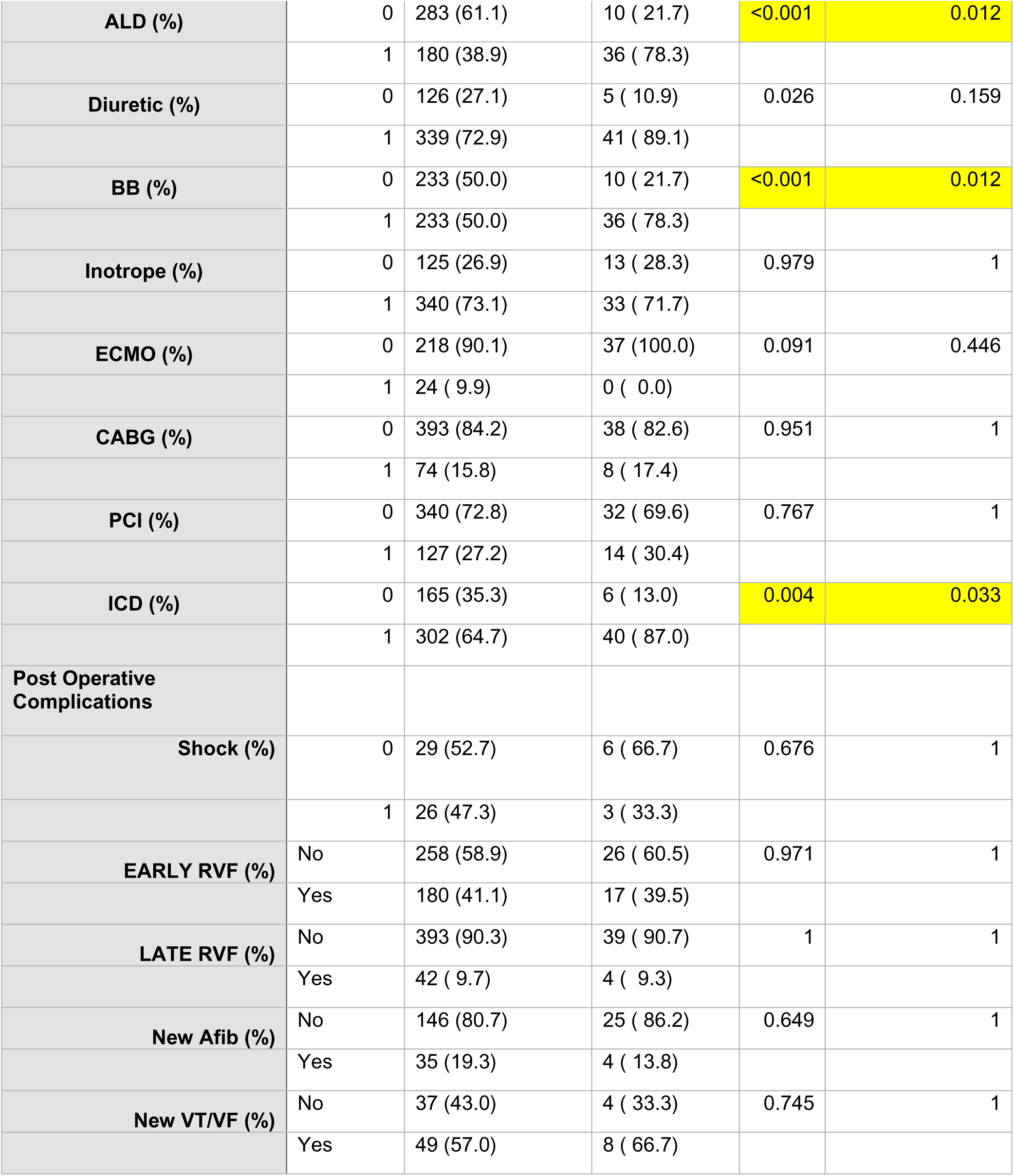
Baseline LVAD Recipient Demographics. Values are presented as median ± interquartile range (IQR), mean ± standard deviation (SD), or number (percentage) for categorical variables. Abbreviations: BMI, body mass index; COPD, chronic obstructive pulmonary disease; DM, diabetes mellittus; MI, myocardial infarction; CVA, cerebrovascular accident; CAD, coronary artery disease; ALD, aldosterone; BB, beta blocker; ECMO, extracorporeal membrane oxygenation; CABG, coronary artery bypass graft; PCI, percutaneous coronary intervention; ICD, implantable cardiac defibrillator; RVF, right ventricular failure; Afib, atrial fibrillation; VT/VF, ventricular tachycardia/ventricular fibrillation.

| Baseline Characteristics |  |  |  |  |  |
| --- | --- | --- | --- | --- | --- |
| Variable |  | No OSA( N=468) | OSA (N=46) | P-value | Adjusted P-value |
| Age (mean (SD)) |  | 54.07 (13.89) | 52.67 (14.00) | 0.516 | 1 |
| Sex (%) | Female | 129 (27.6) | 10 ( 21.7) | 0.5 | 1 |
|  | Male | 339 (72.4) | 36 ( 78.3) |  |  |
| BMI (mean (SD)) |  | 27.04 (5.93) | 31.48 (6.39) | <0.001 | 0.012 |
| Severity (%) | Mild | 0 ( NaN) | 10 ( 21.7) | NaN | NaN |
|  | Moderate | 0 ( NaN) | 17 ( 37.0) |  |  |
|  | Severe | 0 ( NaN) | 19 ( 41.3) |  |  |
| LVAD_Type (%) | HM/HMII | 343 (73.3) | 31 ( 67.4) | 0.439 | 1 |
|  | HM3 | 32 ( 6.8) | 4 ( 8.7) |  |  |
|  | HW/HVAD | 91 (19.4) | 10 ( 21.7) |  |  |
|  | TAH | 2 ( 0.4) | 1 ( 2.2) |  |  |
| Comorbidities |  |  |  |  |  |
| HTN (%) | 0 | 183 (39.2) | 7 ( 15.2) | 0.002 | 0.02 |
|  | 1 | 284 (60.8) | 39 ( 84.8) |  |  |
| COPD (%) | 0 | 417 (89.5) | 38 ( 82.6) | 0.242 | 0.988 |
|  | 1 | 49 (10.5) | 8 ( 17.4) |  |  |
| DM (%) | 0 | 277 (59.2) | 23 ( 50.0) | 0.294 | 1 |
|  | 1 | 191 (40.8) | 23 ( 50.0) |  |  |
| MI (%) | 0 | 297 (63.5) | 26 ( 56.5) | 0.442 | 1 |
|  | 1 | 171 (36.5) | 20 ( 43.5) |  |  |
| CVA (%) | 0 | 407 (87.0) | 42 ( 91.3) | 0.54 | 1 |
|  | 1 | 61 (13.0) | 4 ( 8.7) |  |  |
| CAD (%) | 0 | 214 (45.7) | 23 ( 50.0) | 0.689 | 1 |
|  | 1 | 254 (54.3) | 23 ( 50.0) |  |  |
| ACE (%) | 0 | 377 (81.1) | 33 ( 71.7) | 0.33 | 1 |
|  | 1 | 88 (18.9) | 12 ( 26.1) |  |  |
| ARB (%) | 0 | 409 (87.8) | 31 ( 67.4) | <0.001 | 0.012 |
|  | 1 | 57 (12.2) | 15 ( 32.6) |  |  |

|  |  |  |  |  |  |  |
| --- | --- | --- | --- | --- | --- | --- |
| **ALD (%)** |  | 0 | 283 (61.1) | 10 ( 21.7) | <0.001 | 0.012 |
|  |  | 1 | 180 (38.9) | 36 ( 78.3) |  |  |
| **Diuretic (%)** |  | 0 | 126 (27.1) | 5 ( 10.9) | 0.026 | 0.159 |
|  |  | 1 | 339 (72.9) | 41 ( 89.1) |  |  |
| **BB (%)** |  | 0 | 233 (50.0) | 10 ( 21.7) | <0.001 | 0.012 |
|  |  | 1 | 233 (50.0) | 36 ( 78.3) |  |  |
| **Inotrope (%)** |  | 0 | 125 (26.9) | 13 ( 28.3) | 0.979 | 1 |
|  |  | 1 | 340 (73.1) | 33 ( 71.7) |  |  |
| **ECMO (%)** |  | 0 | 218 (90.1) | 37 (100.0) | 0.091 | 0.446 |
|  |  | 1 | 24 ( 9.9) | 0 ( 0.0) |  |  |
| **CABG (%)** |  | 0 | 393 (84.2) | 38 ( 82.6) | 0.951 | 1 |
|  |  | 1 | 74 (15.8) | 8 ( 17.4) |  |  |
| **PCI (%)** |  | 0 | 340 (72.8) | 32 ( 69.6) | 0.767 | 1 |
|  |  | 1 | 127 (27.2) | 14 ( 30.4) |  |  |
| **ICD (%)** |  | 0 | 165 (35.3) | 6 ( 13.0) | 0.004 | 0.033 |
|  |  | 1 | 302 (64.7) | 40 ( 87.0) |  |  |
| **Post Operative Complications** |  |  |  |  |  |  |
| **Shock (%)** |  | 0 | 29 (52.7) | 6 ( 66.7) | 0.676 | 1 |
|  |  | 1 | 26 (47.3) | 3 ( 33.3) |  |  |
| **EARLY RVF (%)** | No |  | 258 (58.9) | 26 ( 60.5) | 0.971 | 1 |
|  | Yes |  | 180 (41.1) | 17 ( 39.5) |  |  |
| **LATE RVF (%)** | No |  | 393 (90.3) | 39 ( 90.7) | 1 | 1 |
|  | Yes |  | 42 ( 9.7) | 4 ( 9.3) |  |  |
| **New Afib (%)** | No |  | 146 (80.7) | 25 ( 86.2) | 0.649 | 1 |
|  | Yes |  | 35 (19.3) | 4 ( 13.8) |  |  |
| **New VT/VF (%)** | No |  | 37 (43.0) | 4 ( 33.3) | 0.745 | 1 |
|  | Yes |  | 49 (57.0) | 8 ( 66.7) |  |  |

### Postoperative Outcomes, Biochemical and Hemodynamic Parameters

Postoperative complications including shock, early or late right ventricular failure, *de novo* atrial fibrillation, and ventricular arrhythmias. Biochemical and hemodynamic parameters were largely comparable between groups (**Supplemental Table 1**), with a slightly lower direct bilirubin in the OSA group, which was not significant after correction (p = 0.018, adjusted p = 0.152).

### Survival Analysis

Regarding survival after LVAD implantation, no significant difference in overall survival was observed between OSA and No-OSA LVAD recipients (p=0.33; **Figure 2**). In Cox proportional hazards analysis (**Table 2)**, survival was not significantly different between patients with and without OSA (HR 1.03, 95% CI 0.65–1.62; *p* = 0.897). Worse survival was associated with older age (HR 1.021 per year, 95% CI 1.01–1.03; *p* < 0.001), history of CABG (HR 1.47, 95% CI 1.01–2.12; *p* = 0.042), ACE inhibitor use (HR 1.21, 95% CI 1.01–1.45; *p* = 0.044), and beta-blocker use (HR 1.54, 95% CI 1.14–2.08; *p* = 0.005). These findings indicate that OSA status is not associated with post-LVAD survival after covariate adjustment, whereas several baseline comorbidities and medication use are associated with survival.

**Figure 2.**
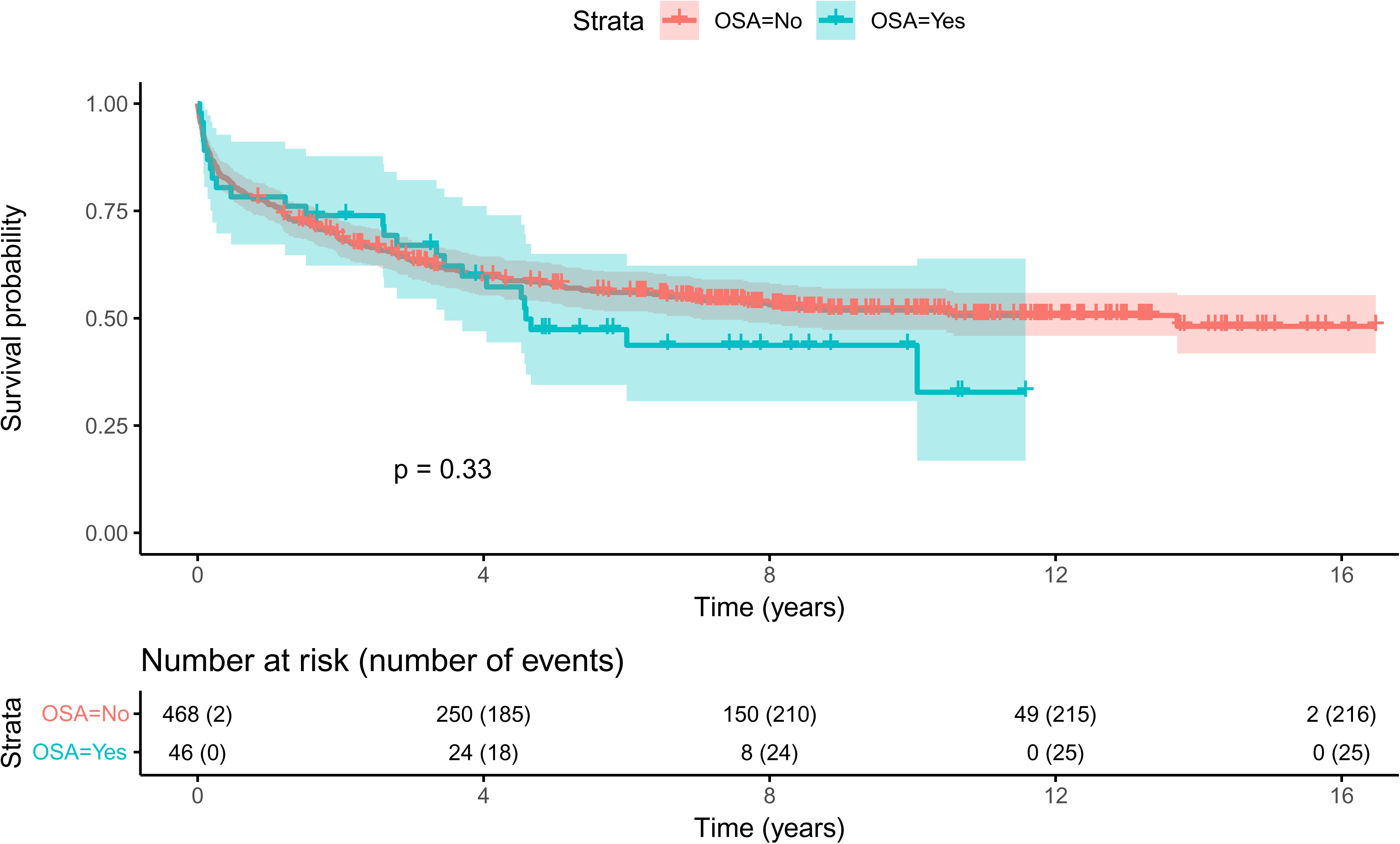
Survival Amongst LVAD Recipients with OSA. Kaplan-Meier survival estimates between LVAD patients with OSA versus No-OSA. 95% confidence intervals are depicted with shading. Patients with OSA had no significant difference in survival compared to No-OSA patients as determined by log-rank testing.

**Table 2:** Multivariate Cox Regression Model Assessing Mortality in All Patients. Cox proportional hazard model for patients with and without OSA after adjusting for other independent predictors of survival. Includes hazard ratios associated with survival following LVAD implantation in patients with OSA.

|  | **Log**<br>**Hazard**<br>**Ratio** | **Hazard**<br>**Ratio** | **95% CI**<br>**(Lower)** | **95% CI**<br>**(Upper)** | **P-value** |
| --- | --- | --- | --- | --- | --- |
| OSA | 0.03 | 1.03 | 0.654 | 1.623 | 0.897 |
| Age | 0.021 | 1.021 | 1.01 | 1.033 | <0.001 |
| Sex (Male vs<br>Female) | 0.173 | 1.189 | 0.869 | 1.627 | 0.278 |
| HTN | 0.198 | 1.219 | 0.907 | 1.639 | 0.189 |
| MI | -0.341 | 0.711 | 0.493 | 1.025 | 0.068 |
| CAD | -0.018 | 0.982 | 0.68 | 1.419 | 0.924 |
| COPD | 0.249 | 1.283 | 0.884 | 1.863 | 0.19 |
| CABG | 0.382 | 1.465 | 1.013 | 2.119 | 0.042 |
| PCI | 0.269 | 1.308 | 0.913 | 1.874 | 0.143 |
| ICD | -0.143 | 0.867 | 0.653 | 1.15 | 0.321 |
| ACE Inhibitor | 0.188 | 1.206 | 1.005 | 1.448 | 0.044 |
| ARB | 0.069 | 1.072 | 0.707 | 1.625 | 0.744 |
| BB | 0.434 | 1.544 | 1.144 | 2.084 | 0.005 |
| ALD | 0.062 | 1.064 | 0.799 | 1.418 | 0.67 |

### CPAP/BiPAP Compliance in OSA Patients

Regarding the 46 LVAD patients with OSA, 24 were compliant with CPAP/BiPAP therapy and 22 were noncompliant (**Table 3**). Compliant patients were slightly younger (48.18 ± 15.89 vs. 58.96 ± 10.32 years; p = 0.01), though this difference was not statistically significant after correction (p = 0.07). There were no significant differences in sex, BMI, LVAD type, OSA severity, or baseline comorbidities (**Table 3**). Postoperative complications were also similar between groups. While biochemical profiles were comparable, hemodynamic assessment showed trends toward higher right atrial pressure (RAP), pulmonary artery pressures, and pulmonary capillary wedge pressure (PCWP) in compliant patients, but these differences did not reach significance after adjustment (**Supplemental Table 2**). Notably, pulmonary vascular resistance by thermodilution (PVR TD) was lower in compliant patients (−5.52 ± 2.17 vs. −0.91 ± 1.94; unadjusted p = 0.031), suggesting improved pulmonary hemodynamics, although this was not statistically significant after correction.

**Table 3:**
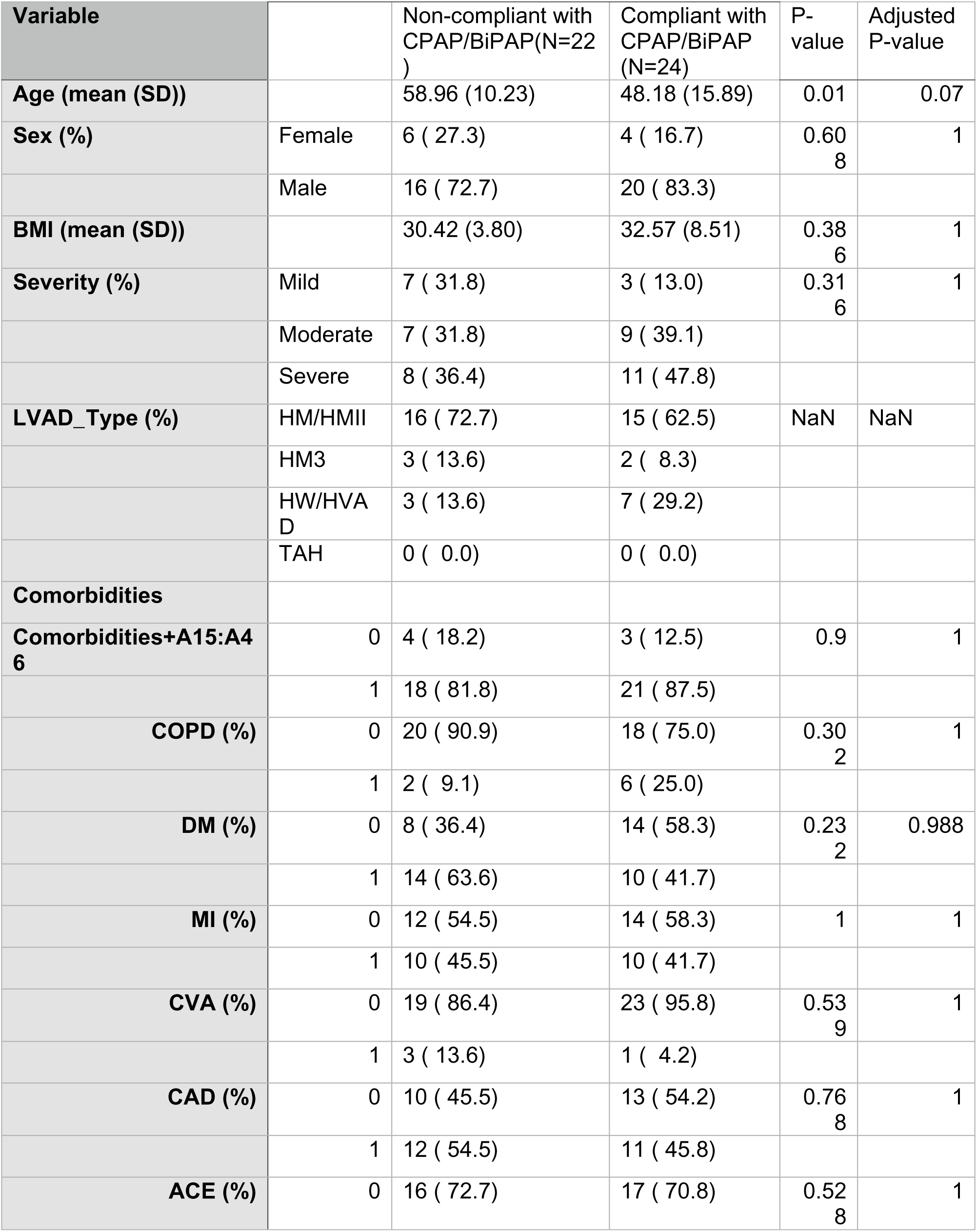
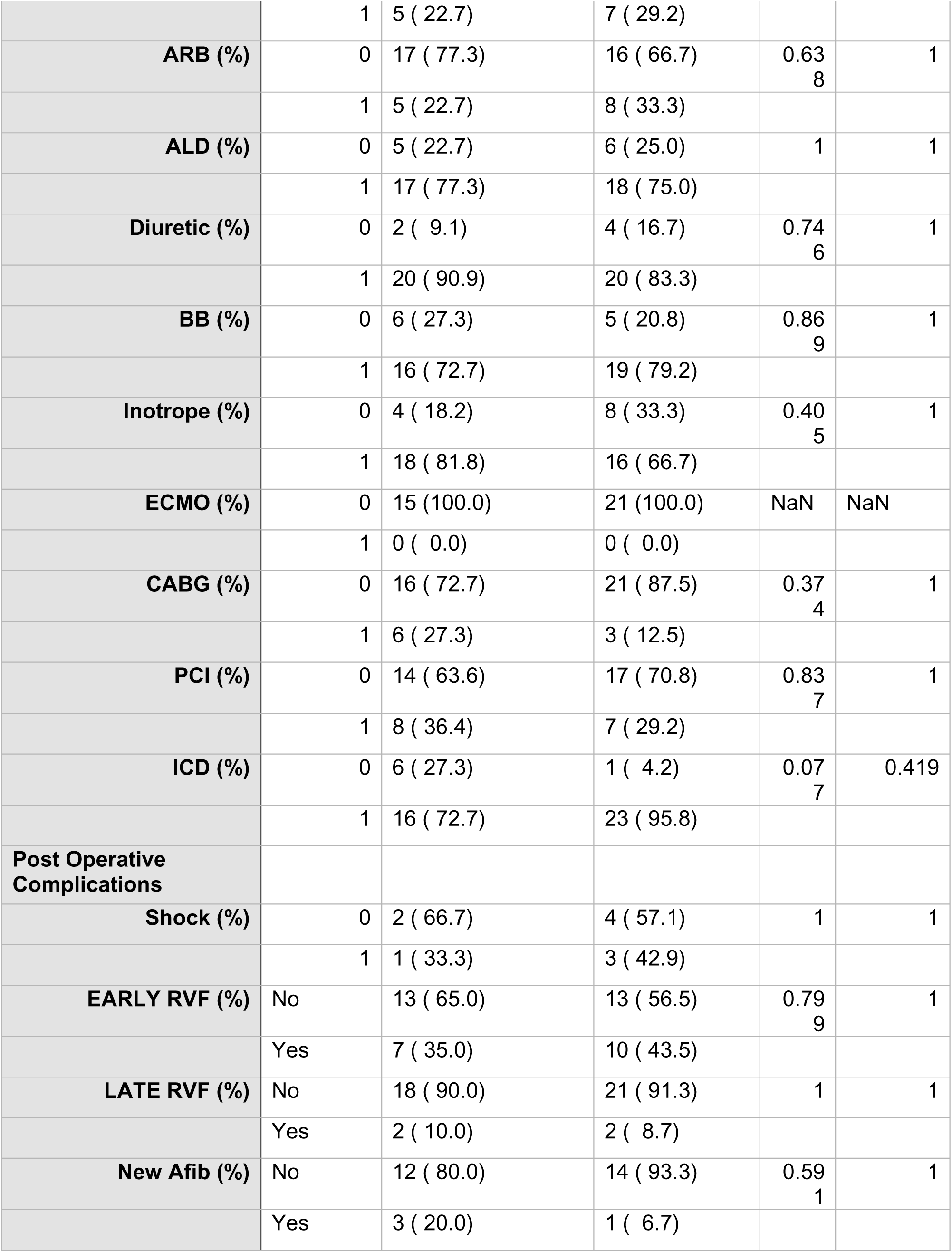
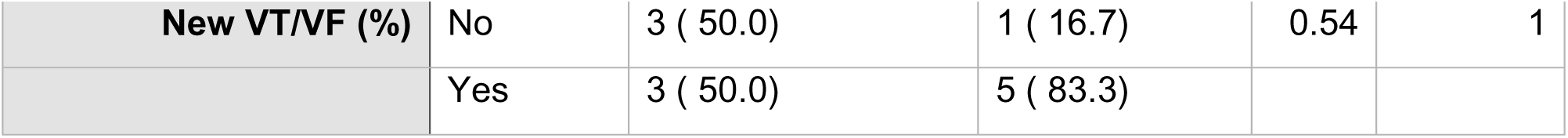
Baseline CPAP/BiPAP Characteristics. Values are presented as median ± interquartile range (IQR), mean ± standard deviation (SD), or number (percentage) for categorical variables. Abbreviations: BMI, body mass index; COPD, chronic obstructive pulmonary disease; DM, diabetes mellittus; MI, myocardial infarction; CVA, cerebrovascular accident; CAD, coronary artery disease; ALD, aldosterone; BB, beta blocker; ECMO, extracorporeal membrane oxygenation; CABG, coronary artery bypass graft; PCI, percutaneous coronary intervention; ICD, implantable cardiac defibrillator; RVF, right ventricular failure; Afib, atrial fibrillation; VT/VF, ventricular tachycardia/ventricular fibrillation;

| Variable |  | Non-compliant with CPAP/BiPAP(N=22 ) | Compliant with CPAP/BiPAP (N=24) | P-value | Adjusted P-value |
| --- | --- | --- | --- | --- | --- |
| **Age (mean (SD))** |  | 58.96 (10.23) | 48.18 (15.89) | 0.01 | 0.07 |
| **Sex (%)** | Female | 6 ( 27.3) | 4 ( 16.7) | 0.608 | 1 |
|  | Male | 16 ( 72.7) | 20 ( 83.3) |  |  |
| **BMI (mean (SD))** |  | 30.42 (3.80) | 32.57 (8.51) | 0.386 | 1 |
| **Severity (%)** | Mild | 7 ( 31.8) | 3 ( 13.0) | 0.316 | 1 |
|  | Moderate | 7 ( 31.8) | 9 ( 39.1) |  |  |
|  | Severe | 8 ( 36.4) | 11 ( 47.8) |  |  |
| **LVAD_Type (%)** | HM/HMII | 16 ( 72.7) | 15 ( 62.5) | NaN | NaN |
|  | HM3 | 3 ( 13.6) | 2 ( 8.3) |  |  |
|  | HW/HVAD | 3 ( 13.6) | 7 ( 29.2) |  |  |
|  | TAH | 0 ( 0.0) | 0 ( 0.0) |  |  |
| **Comorbidities** |  |  |  |  |  |
| **Comorbidities+A15:A46** | 0 | 4 ( 18.2) | 3 ( 12.5) | 0.9 | 1 |
|  | 1 | 18 ( 81.8) | 21 ( 87.5) |  |  |
| **COPD (%)** | 0 | 20 ( 90.9) | 18 ( 75.0) | 0.302 | 1 |
|  | 1 | 2 ( 9.1) | 6 ( 25.0) |  |  |
| **DM (%)** | 0 | 8 ( 36.4) | 14 ( 58.3) | 0.232 | 0.988 |
|  | 1 | 14 ( 63.6) | 10 ( 41.7) |  |  |
| **MI (%)** | 0 | 12 ( 54.5) | 14 ( 58.3) | 1 | 1 |
|  | 1 | 10 ( 45.5) | 10 ( 41.7) |  |  |
| **CVA (%)** | 0 | 19 ( 86.4) | 23 ( 95.8) | 0.539 | 1 |
|  | 1 | 3 ( 13.6) | 1 ( 4.2) |  |  |
| **CAD (%)** | 0 | 10 ( 45.5) | 13 ( 54.2) | 0.768 | 1 |
|  | 1 | 12 ( 54.5) | 11 ( 45.8) |  |  |
| **ACE (%)** | 0 | 16 ( 72.7) | 17 ( 70.8) | 0.528 | 1 |

|  |  |  |  |  |  |  |
| --- | --- | --- | --- | --- | --- | --- |
|  |  | 1 | 5 ( 22.7) | 7 ( 29.2) |  |  |
| **ARB (%)** |  | 0 | 17 ( 77.3) | 16 ( 66.7) | 0.63<br>8 | 1 |
|  |  | 1 | 5 ( 22.7) | 8 ( 33.3) |  |  |
| **ALD (%)** |  | 0 | 5 ( 22.7) | 6 ( 25.0) | 1 | 1 |
|  |  | 1 | 17 ( 77.3) | 18 ( 75.0) |  |  |
| **Diuretic (%)** |  | 0 | 2 ( 9.1) | 4 ( 16.7) | 0.74<br>6 | 1 |
|  |  | 1 | 20 ( 90.9) | 20 ( 83.3) |  |  |
| **BB (%)** |  | 0 | 6 ( 27.3) | 5 ( 20.8) | 0.86<br>9 | 1 |
|  |  | 1 | 16 ( 72.7) | 19 ( 79.2) |  |  |
| **Inotrope (%)** |  | 0 | 4 ( 18.2) | 8 ( 33.3) | 0.40<br>5 | 1 |
|  |  | 1 | 18 ( 81.8) | 16 ( 66.7) |  |  |
| **ECMO (%)** |  | 0 | 15 (100.0) | 21 (100.0) | NaN | NaN |
|  |  | 1 | 0 ( 0.0) | 0 ( 0.0) |  |  |
| **CABG (%)** |  | 0 | 16 ( 72.7) | 21 ( 87.5) | 0.37<br>4 | 1 |
|  |  | 1 | 6 ( 27.3) | 3 ( 12.5) |  |  |
| **PCI (%)** |  | 0 | 14 ( 63.6) | 17 ( 70.8) | 0.83<br>7 | 1 |
|  |  | 1 | 8 ( 36.4) | 7 ( 29.2) |  |  |
| **ICD (%)** |  | 0 | 6 ( 27.3) | 1 ( 4.2) | 0.07<br>7 | 0.419 |
|  |  | 1 | 16 ( 72.7) | 23 ( 95.8) |  |  |
| **Post Operative Complications** |  |  |  |  |  |  |
| **Shock (%)** |  | 0 | 2 ( 66.7) | 4 ( 57.1) | 1 | 1 |
|  |  | 1 | 1 ( 33.3) | 3 ( 42.9) |  |  |
| **EARLY RVF (%)** | No |  | 13 ( 65.0) | 13 ( 56.5) | 0.79<br>9 | 1 |
|  | Yes |  | 7 ( 35.0) | 10 ( 43.5) |  |  |
| **LATE RVF (%)** | No |  | 18 ( 90.0) | 21 ( 91.3) | 1 | 1 |
|  | Yes |  | 2 ( 10.0) | 2 ( 8.7) |  |  |
| **New Afib (%)** | No |  | 12 ( 80.0) | 14 ( 93.3) | 0.59<br>1 | 1 |
|  | Yes |  | 3 ( 20.0) | 1 ( 6.7) |  |  |
| New VT/VF (%) | No | 3 ( 50.0) | 1 ( 16.7) | 0.54 | 1 |
|  | Yes | 3 ( 50.0) | 5 ( 83.3) |  |  |

No significant difference in survival was observed among patients with mild, moderate, and severe OSA (**Supplemental Figure 1**). However, a significant difference in survival was observed between CPAP/BiPAP-compliant patients. CPAP/BiPAP-compliant patients had significantly better survival compared to noncompliant OSA patients (p=0.0099; **Figure 3**)

**Figure 3.**
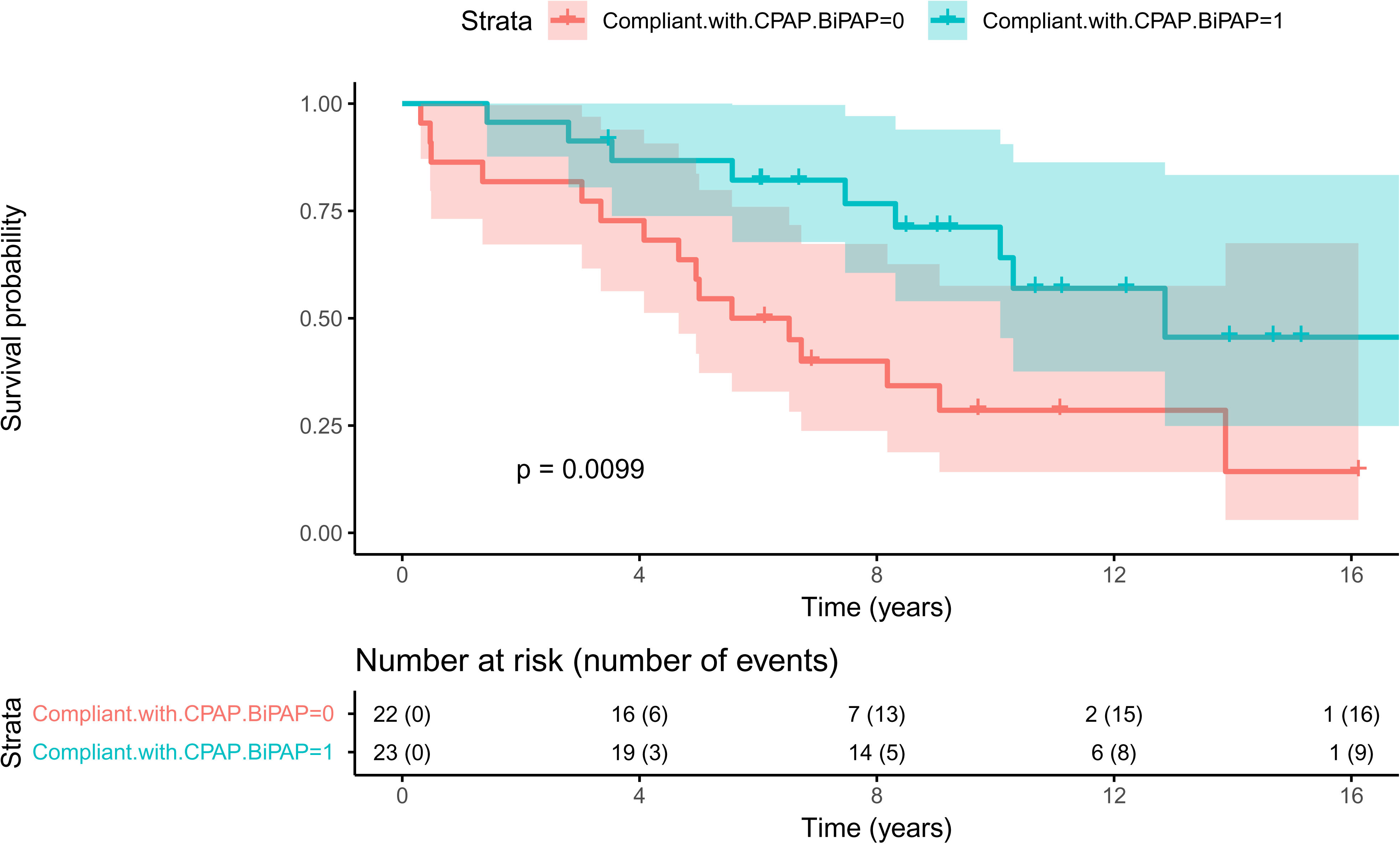
Survival Amongst CPAP/BiPAP Compliant LVAD Recipients with OSA. Kaplan-Meier survival estimates for OSA patients compliant with CPAP/BiPAP versus noncompliant with CPAP/BiPAP. 95% confidence intervals are depicted with shading. CPAP/BiPAP noncompliant patients had a significantly lower probability of survival than compliant patients.

## DISCUSSION

OSA has been shown to lead to worse outcomes in the HF population; however, its prevalence and impact in patients with newly implanted LVADs remain poorly defined. Prior work by Kumai et. al reported that 24% of LVAD patients suffer from sleep-disordered breathing. In comparison, OSA prevalence in the general HF population exceeds 60% (4). In our single-center cohort, OSA was identified in 8.9% of newly implanted LVAD patients, a rate similar to previously reported values (15).

In terms of survival, LVAD patients with OSA had outcomes comparable to those without OSA **(Figure 2**). Multivariate analysis identified that prior cardiovascular dysfunction, rather than OSA itself as an independent predictor of LVAD survival. Interestingly, a significant survival difference emerged when stratifying OSA patients by CPAP/BiPAP compliance. Patients who were adherent to CPAP/BiPAP demonstrated improved survival.

### Prevalence and Under-Diagnosis

In the general heart failure population, OSA prevalence is estimated to exceed 60% (4). In contrast, our retrospective cohort identified a pre-implant OSA diagnosis in only 8.9% of patients. This discrepancy likely reflects significant under-diagnosis in the clinical setting rather than a true biological difference in our cohort. Consequently, our “No-OSA” control group likely contained a substantial number of patients with undiagnosed, untreated sleep apnea. This selection bias in the control group may explain why we observed no statistical difference in survival between the broad “OSA” and “No-OSA” cohorts. However, the survival divergence observed within the diagnosed group, stratified by compliance, remains a robust finding that underscores the severity of untreated sleep-disordered breathing.

### Mechanistic Considerations

The survival benefit observed in compliant patients is likely driven by favorable hemodynamic modification. In non-compliant patients, apneic events generate negative intrathoracic pressure, increasing venous return (preload), while hypoxia and hypercapnia simultaneously induce pulmonary vasoconstriction and increase right ventricular (RV) afterload. This mismatch can precipitate RV dilation, interventricular septal bowing, and impaired LV filling, which are particularly deleterious in LVAD physiology.

LVADs are preload-dependent; therefore, RV failure directly compromises LVAD flow. By preventing airway collapse, CPAP/BiPAP stabilizes intrathoracic pressure and reduces pulmonary vascular resistance (PVR). Our data suggests that compliant patients exhibited more favorable pulmonary hemodynamics, potentially protecting the RV from the dual insult of operative stun and apneic stress. This synergistic protection likely accounts for the survival advantage seen in the compliant cohort.

### Device-Specific Considerations

The interplay between OSA and device physiology is complex. While our study spanned an era including pulsatile-flow (HM1) devices, modern continuous-flow (CF) pumps (HMII, HM3) dominate current practice. CF-LVADs are less sensitive to preload changes than native ventricles but are prone to “suction events” if RV failure leads to under-filling of the left ventricle. Apnea-induced RV dysfunction can therefore cause unpredictable drops in LVAD flow and trigger low-flow alarms or arrhythmias (18). CPAP therapy essentially “stents” the airway and the pulmonary vasculature, ensuring consistent LV preloading and allowing continuous-flow devices to operate at optimal parameters without suction or speed drops.

### Clinical Implications

Current guidelines for LVAD evaluation are rigorous, yet standardized screening for sleep-disordered breathing remains variable. Our findings indicate that NIPPV is a high-yield, low-risk intervention. The survival divergence suggests that “diagnosed and treated” OSA carries a better prognosis than “diagnosed and untreated” OSA. Given the high prevalence of undiagnosed OSA in heart failure, it is reasonable to advocate for aggressive PSG screening in all LVAD candidates. Early initiation of therapy, even in the bridge-to-transplant or destination-therapy workup phase, may precondition the RV for the hemodynamic rigors of post-implantation support. These findings align with prior observational studies showing CPAP therapy in patients with HF and OSA improves hemodynamics and may reduce mortality and rehospitalization (2,10).

### Study Limitations

This study is limited by its single-center, retrospective design and the reliance on provider documentation for compliance rather than device downloads, which may introduce classification bias. Additionally, the low diagnostic rate (8.9%) suggests selection bias, in which only the most symptomatic patients were screened. Finally, because our cohort spans 15 years, it includes older generation devices; however, the physiologic argument for RV protection remains relevant for current centrifugal-flow pumps. Finally, OSA severity was determined by AHI, which may not accurately reflect hypoxic burden or differentiate between REM and non-REM events, limiting the assessment of OSA severity on outcomes.

## CONCLUSION

In patients with LVADs, CPAP/BiPAP adherence is a significant determinant of long-term survival. The survival advantage associated with compliance suggests that the deleterious effects of OSA are reversible. These data support a paradigm shift toward mandatory OSA screening and aggressive management of sleep-disordered breathing as standard components of multidisciplinary LVAD care.

## Data Availability

The authors declare that all supporting data are available within the article and its online supplementary files.

## Acknowledgements and funding sources

This work was supported by T32HL134616 to S.S.S.; A generous donation by Bob and Corrine Frick (Bob Frick Research Chair in Heart Failure and Arrhythmia for S.A.S.); Dept of IM Pilot Award for 2023-2024 and NIH R01 HL160590; and R01 NIHMS1018036 to S.A.S.

## Author contributions

JC, SSS, and SAS wrote the original manuscript. JC and SSS collected data. SSS, MJ, JP performed formal analysis of the data. SSS and SAS determine the experimental design and rigor. JC, SSS, JW, MJ, JP, NAM, BAW, and SAS validated the results. All authors edited and approved the final version of the manuscript.

## LIST OF ABBREVIATIONS

BiPAP: Bilevel positive airway pressure
CPAP: Continuous positive airway pressure
HF: Heart failure
LVAD: Left ventricular assist device
NIPPV: Noninvasive positive pressure ventilation
OSA: Obstructive sleep apnea

---

**Supplemental Figure 1:**
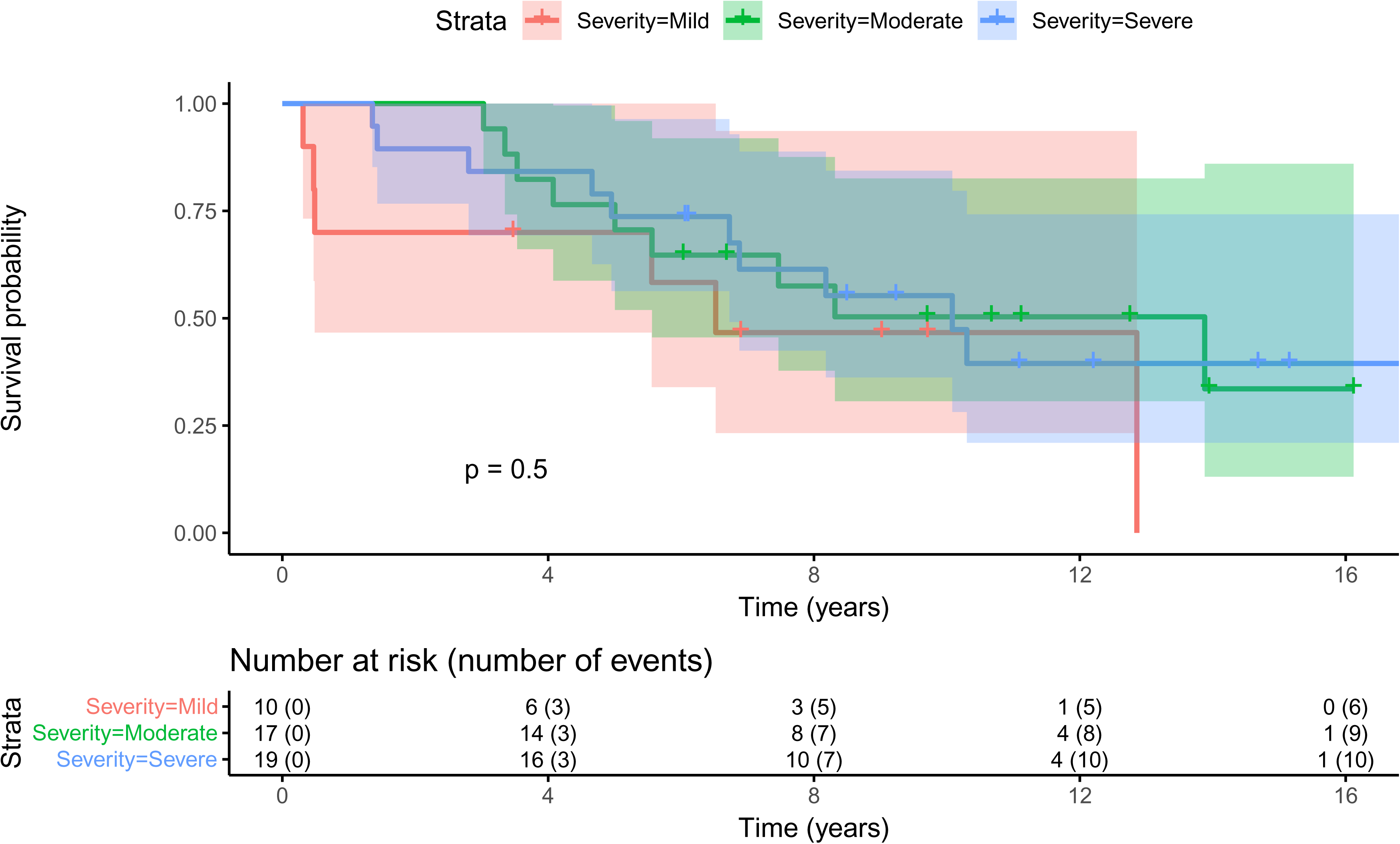
Impact of OSA Severity on Survival. Kaplan-Meier survival estimates for OSA patients based on degree of severity. 95% confidence intervals are depicted with shading. No difference in survival was observed.

**Supplemental Table 1:**
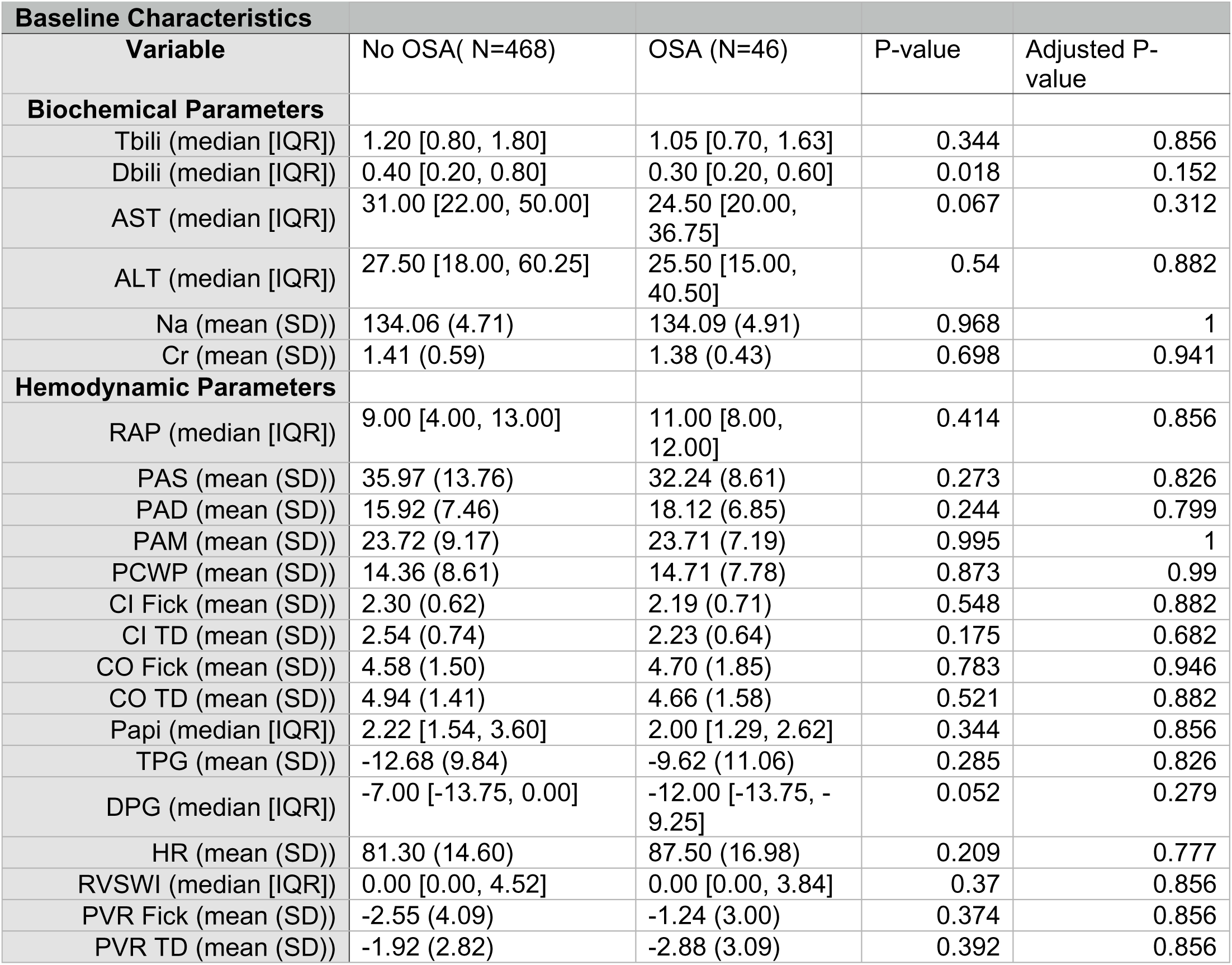
Baseline Biochemical and Hemodynamic Parameters in LVAD recipients with OSA. Values are presented as median ± interquartile range (IQR), mean ± standard deviation (SD), or number (percentage) for categorical variables. Abbreviations: Tbili, total bilirubin; Dbili, direct bilirubin; Na, sodium; Cr, creatinine; RAP, right atrial pressure; PAS, pulmonary artery systolic pressure; PAD, pulmonary artery diastolic pressure; PAM, pulmonary artery mean pressure; PCWP, pulmonary capillary wedge pressure; CI, cardiac index; CO, cardiac output; Papi, pulmonary artery pulsatility index; TPG, transpulmonary pressure gradient; DPG, diastolic pulmonary gradient; HR, heart rate; RVSWI, right ventricular stroke work index; PVR, pulmonary vascular resistance.

**Supplemental Table 2:** Baseline Biochemical and Hemodynamic Parameters Based on CPAP/BiPAP Compliance. Values are presented as median ± interquartile range (IQR), mean ± standard deviation (SD), or number (percentage) for categorical variables. Abbreviations: Tbili, total bilirubin; Dbili, direct bilirubin; Na, sodium; Cr, creatinine; RAP, right atrial pressure; PAS, pulmonary artery systolic pressure; PAD, pulmonary artery diastolic pressure; PAM, pulmonary artery mean pressure; PCWP, pulmonary capillary wedge pressure; CI, cardiac index; CO, cardiac output; Papi, pulmonary artery pulsatility index; TPG, transpulmonary pressure gradient; DPG, diastolic pulmonary gradient; HR, heart rate; RVSWI, right ventricular stroke work index; PVR, pulmonary vascular resistance.

| Baseline Characteristics |  |  |  |  |
| --- | --- | --- | --- | --- |
| Variable | Non-compliant with CPAP/BiPAP(N=22) | Compliant with CPAP/BiPAP (N=24) | P-value | Adjusted P-value |
| **Biochemical Parameters** |  |  |  |  |
| Tbili (mean (SD)) | 1.18 (0.59) | 1.48 (1.12) | 0.285 | 0.826 |
| Dbili (mean (SD)) | 0.38 (0.28) | 0.55 (0.63) | 0.249 | 0.799 |
| AST (mean (SD)) | 39.90 (34.30) | 81.04 (236.29) | 0.434 | 0.856 |
| ALT (mean (SD)) | 77.95 (171.24) | 89.43 (239.45) | 0.857 | 0.99 |
| Na (mean (SD)) | 134.41 (3.75) | 133.75 (5.91) | 0.657 | 0.941 |
| Cr (mean (SD)) | 1.53 (0.41) | 1.29 (0.42) | 0.054 | 0.279 |
| **Hemodynamic Parameters** |  |  |  |  |
| RAP (mean (SD)) | 6.50 (3.66) | 13.78 (5.07) | 0.004 | 0.047 |
| PAS (mean (SD)) | 28.00 (8.07) | 36.00 (7.57) | 0.052 | 0.279 |
| PAD (mean (SD)) | 14.38 (4.98) | 21.44 (6.75) | 0.028 | 0.186 |
| PAM (mean (SD)) | 19.62 (5.63) | 27.33 (6.65) | 0.022 | 0.171 |
| PCWP (mean (SD)) | 10.12 (3.00) | 18.78 (8.57) | 0.016 | 0.149 |
| CI Fick (mean (SD)) | 2.08 (0.49) | 2.26 (0.83) | 0.677 | 0.941 |
| CI TD (mean (SD)) | 2.31 (0.38) | 2.17 (0.80) | 0.739 | 0.941 |
| CO Fick (mean (SD)) | 4.32 (1.34) | 4.90 (2.12) | 0.594 | 0.921 |
| CO TD (mean (SD)) | 4.43 (1.07) | 4.82 (1.93) | 0.696 | 0.941 |
| Papi (mean (SD)) | 3.47 (3.03) | 1.17 (0.68) | 0.176 | 0.682 |
| TPG (mean (SD)) | -7.62 (7.35) | -12.80 (15.87) | 0.436 | 0.856 |
| DPG (mean (SD)) | -10.00 (3.21) | -14.67 (2.52) | 0.058 | 0.284 |
| HR (mean (SD)) | 84.67 (15.17) | 91.75 (21.00) | 0.55 | 0.882 |
| RVSWI (mean (SD)) | 1.63 (2.42) | 2.16 (2.83) | 0.769 | 0.941 |
| PVR Fick (mean (SD)) | -0.88 (1.61) | -1.60 (4.25) | 0.76 | 0.941 |
| PVR TD (mean (SD)) | -0.91 (1.94) | -5.52 (2.17) | 0.031 | 0.192 |

## REFERENCES

1. Sharma B, Owens R, Malhotra A. Sleep in congestive heart failure. Med Clin North Am. 2010 May;94(3):447–64.

2. Wang H, Parker JD, Newton GE, Floras JS, Mak S, Chiu KL, et al. Influence of obstructive sleep apnea on mortality in patients with heart failure. J Am Coll Cardiol. 2007 Apr 17;49(15):1625–31.

3. Bradley TD, Hall MJ, Ando S, Floras JS. Hemodynamic effects of simulated obstructive apneas in humans with and without heart failure. Chest. 2001 Jun;119(6):1827–35.

4. Javaheri S, Javaheri S. Obstructive Sleep Apnea in Heart Failure: Current Knowledge and Future Directions. J Clin Med. 2022 Jun 16;11(12):3458.

5. Akkanti B, Castriotta RJ, Sayana P, Nunez E, Rajapreyar I, Kumar S, et al. Ventricular assist devices and sleep-disordered breathing. Sleep Med Rev. 2017 Oct;35:51–61.

6. Kumai Y, Seguchi O, Mochizuki H, Kimura Y, Iwasaki K, Kuroda K, et al. Impact of sleep-disordered breathing on ventricular tachyarrhythmias after left ventricular assist device implantation. J Artif Organs Off J Jpn Soc Artif Organs. 2022 Sep;25(3):223–30.

7. Vazir A, Hastings PC, Morrell MJ, Pepper J, Henein MY, Westaby S, et al. Resolution of central sleep apnoea following implantation of a left ventricular assist device. Int J Cardiol. 2010 Feb 4;138(3):317–9.

8. Vermes E, Fonkoua H, Kirsch M, Damy T, Margarit L, Hillion ML, et al. Resolution of sleep-disordered breathing with a biventricular assist device and recurrence after heart transplantation. J Clin Sleep Med JCSM Off Publ Am Acad Sleep Med. 2009 Jun 15;5(3):248–50.

9. Yeghiazarians Y, Jneid H, Tietjens JR, Redline S, Brown DL, El-Sherif N, et al. Obstructive Sleep Apnea and Cardiovascular Disease: A Scientific Statement From the American Heart Association. Circulation. 2021 Jul 20;144(3):e56–67.

10. Kasai T, Narui K, Dohi T, Yanagisawa N, Ishiwata S, Ohno M, et al. Prognosis of patients with heart failure and obstructive sleep apnea treated with continuous positive airway pressure. Chest. 2008 Mar;133(3):690–6.

11. Kaneko Y, Floras JS, Usui K, Plante J, Tkacova R, Kubo T, et al. Cardiovascular effects of continuous positive airway pressure in patients with heart failure and obstructive sleep apnea. N Engl J Med. 2003 Mar 27;348(13):1233–41.

12. Mansfield DR, Gollogly NC, Kaye DM, Richardson M, Bergin P, Naughton MT. Controlled trial of continuous positive airway pressure in obstructive sleep apnea and heart failure. Am J Respir Crit Care Med. 2004 Feb 1;169(3):361–6.

13. McEvoy RD, Antic NA, Heeley E, Luo Y, Ou Q, Zhang X, et al. CPAP for Prevention of Cardiovascular Events in Obstructive Sleep Apnea. N Engl J Med. 2016 Sep 8;375(10):919–31.

14. Soliman OII, Akin S, Muslem R, Boersma E, Manintveld OC, Krabatsch T, et al. Derivation and Validation of a Novel Right-Sided Heart Failure Model After Implantation of Continuous Flow Left Ventricular Assist Devices. Circulation. 2018 Feb 27;137(9):891–906.

15. Cowie MR, Gallagher AM. Sleep Disordered Breathing and Heart Failure: What Does the Future Hold? JACC Heart Fail. 2017 Oct;5(10):715–23.

16. Cheng A, Williamitis CA, Slaughter MS. Comparison of continuous-flow and pulsatile-flow left ventricular assist devices: is there an advantage to pulsatility? Ann Cardiothorac Surg. 2014 Nov;3(6):573–81.

17. Laoutaris ID. Restoring pulsatility and peakVO2 in the era of continuous flow, fixed pump speed, left ventricular assist devices: “A hypothesis of pump’s or patient’s speed?” Eur J Prev Cardiol. 2019 Nov;26(17):1806–15.

18. Lim HS, Howell N, Ranasinghe A. The Physiology of Continuous-Flow Left Ventricular Assist Devices. J Card Fail. 2017 Feb;23(2):169–80.

19. Fukamachi K, Shiose A, Massiello A, Horvath DJ, Golding LAR, Lee S, et al. Preload Sensitivity in Cardiac Assist Devices. Ann Thorac Surg. 2013 Jan;95(1):373–80.

20. Letsou GV, Pate TD, Gohean JR, Kurusz M, Longoria RG, Kaiser L, et al. Improved left ventricular unloading and circulatory support with synchronized pulsatile left ventricular assistance compared with continuous-flow left ventricular assistance in an acute porcine left ventricular failure model. J Thorac Cardiovasc Surg. 2010 Nov;140(5):1181–8.

21. Kendzerska T, Gershon AS, Hawker G, Leung RS, Tomlinson G. Obstructive sleep apnea and risk of cardiovascular events and all-cause mortality: a decade-long historical cohort study. PLoS Med. 2014 Feb;11(2):e1001599.

22. Soori R, Baikunje N, D’sa I, Bhushan N, Nagabhushana B, Hosmane GB. Pitfalls of AHI system of severity grading in obstructive sleep apnoea. Sleep Sci Sao Paulo Braz. 2022;15(Spec 1):285–8.

23. Zhang L, Wu H, Zhang X, Wei X, Hou F, Ma Y. Sleep heart rate variability assists the automatic prediction of long-term cardiovascular outcomes. Sleep Med. 2020 Mar;67:217–24.

24. Shamim-Uzzaman QA, Singh S, Chowdhuri S. Hypopnea definitions, determinants and dilemmas: a focused review. Sleep Sci Pract. 2018 May 23;2(1):7.

